# Empirically grounded projections of shifts in 24-hour movement behaviours under climate change–driven warming

**DOI:** 10.64898/2026.02.08.26345870

**Authors:** Ty Ferguson, Carol Maher, Rachel Curtis, Francois Fraysse, Bastien Lechat, Suzanne Mavoa, Sebastien Chastin

**Author notes:** **Corresponding author** Ty Ferguson.

## Abstract

**Introduction:** Climate change is expected to alter daily patterns of sleep, sedentary behaviour and physical activity, yet empirically grounded projections across the full 24-hour movement spectrum are lacking. This study estimated how projected future warming may alter 24-hour movement behaviour patterns in adults.

**Methods:** A Monte Carlo simulation framework estimated temperature-dependent distributions of daily movement behaviour duration using data from 368 adults in the Annual Rhythms in Adults (ARIA) study in Adelaide, Australia. A total of 85,182 valid person-days were linked to daily temperature data to determine empirical temperature-behaviour relationships. The resulting distributions were used to simulate behaviour under five Intergovernmental Panel on Climate Change warming scenarios (+1.5°C to +4.4°C above pre-industrial levels) across a full calendar year relative to current-climate conditions (+0.99°C above pre-industrial levels).

**Results:** Simulations projected small but consistent behavioural shifts with warming. Annual median increased for MVPA (+49min to +4h 22min per person) and LPA (+3h to +13h 1min per person), while sleep declined (–5h 29min to –23h 19min per person). Physical activity gains were concentrated in cooler months, whereas sleep losses persisted year-round. Changes in sedentary behaviour were minimal and inconsistent.

**Discussion:** Rising temperatures may modestly increase year-long physical activity but reduce sleep duration, in a temperate-zone Mediterranean climate geography producing meaningful cumulative health implications. However, these might be confounded by the effect of other meteorological changes such as rainfall and humidity, which warrant further investigation.

## Introduction

Climate change is one of the greatest existential threats of this century.^1^ The world is experiencing shifts in weather patterns: rising temperatures, altered rainfall, and more frequent extreme weather events.^2-4^ These changes have wide ranging implications potentially affecting natural and built environments, food security, population displacement, and human health.^5,6^

Environmental changes and their associated weather patterns have significant implications for global human health.^6,7^ For instance, increased exposure to high temperatures has been associated with higher rates of illness and mortality,^8^ more frequent flooding may increase the transmission of communicable diseases,^9^ and wildfires are associated with elevated risk of respiratory symptoms burden.^10^ Critically, the health impacts of climate change are not evenly distributed: vulnerable populations face greater exposure and fewer resources to adapt, potentially exacerbating existing inequities.^11^

While much research has focused on the direct links between environmental factors and health outcomes, less attention has been given to the potential impacts of changing climate on sleep, physical activity and sedentary behaviour patterns. These daily activities are recognised as important determinants of chronic disease,^12^ mental health,^13^ mortality^14^ and preventive measures of infectious diseases^15^ yet their sensitivity to climate change remains underexplored.

Current literature suggests that sleep, sedentary behaviour and physical activity patterns are associated with seasonal cycles and weather conditions.^16^ For example, less physical activity occurs at both high and low extremes of temperature along with increased precipitation, and less sleep as temperature rises.^17,18^ The few studies examined the impact of climate change on sleep^19,20^ explored global datasets, whilst Li et. al.^21^ explored mainland China residents across millions of sleep records. The findings suggest rising temperatures will erode global sleep by up to 50-58 hours per person per year,^19^ sleep insufficiency will increase by 10.5%,^21^ and predict a 45% higher likelihood of obstructive sleep apnoea (OSA) on hot nights, estimated to double the OSA burden by 2100.^20^ We are still missing information about the effects of rising temperature on physical activity and sedentary behaviours. Understanding and being able to project how 24-hour movement behaviour patterns may shift under future climate scenarios is crucial for developing effective public health strategies in the face of climate change.^22^ This necessity was highlighted by a recent Australian government review.^4^ Without this knowledge, it is difficult to plan current and future intervention priorities related to daily activity patterns.

Building on this emerging literature, we extend the focus beyond sleep, to the full 24-hour movement behaviour spectrum, using a unique year-long empirical dataset from a cohort study in a temperate-zone Mediterranean climate. We use minute-by-minute 24-hour activity data, combined with local weather conditions, to build a model estimating the effect of future temperature scenarios on movement behaviours.

## Methods

### Study design

This study was a secondary analysis of data from the Annual Rhythms In Adults’ lifestyle and health (ARIA) study. The ARIA study followed a cohort of Australian adults over a 13-month period, collecting data on body weight, 24-hour movement behaviours, dietary intake, and wellbeing. Full details of the study protocol are available in a previously published protocol.^23^ Ethics approval was granted by the University of South Australia Human Research Ethics Committee (Protocol no. 201901), and the study was prospectively registered with the Australian New Zealand Clinical Trials Registry (ACTRN12619001430123).

### Setting and participants

Participants were recruited from metropolitan Adelaide, South Australia. Adelaide has a temperate-climate^24^ with a population of approximately 1.4 million people.^25^ A total of 375 community-dwelling adults were enrolled in the ARIA study. Recruitment occurred through two main pathways: parents of children participating in the *Life on Holidays* cohort study^26^ (cohorts 1 and 2), and parents of children aged 5–12 years living in Adelaide (cohort 3). Cohort 3 was recruited through general community outreach, including social media, digital advertising, and notice boards.

Participants were enrolled in two waves: cohort 1 began on 1 December 2019, while cohorts 2 and 3 commenced on 1 December 2020. Each cohort remained active for 13 months, concluding on 31 December of the following year (2020 and 2021, respectively). The study was active across 762 calendar days.

Eligibility criteria included being aged 18–65 years, a parent or guardian of a child either in the Life on Holidays study or aged 5–12 years, residing in the Adelaide metropolitan area, having internet access via a Bluetooth-capable phone or computer, fluency in English, and the ability to ambulate independently. Exclusion criteria included pregnancy, use of an implanted electronic medical device, or having a life-threatening health condition that could affect daily functioning.

Participants received a home visit from a researcher 1 to 3 months before starting the study. During this visit, baseline measures were taken, and participants were provided with a Fitbit Charge 3 activity tracker and a Fitbit Aria body weight scale (either Aria 2 or Aria Air; Fitbit Inc., San Francisco, CA, USA). Participants were asked to wear and sync their Fitbit devices daily and complete eight follow-up surveys focused on wellbeing, dietary intake and vacation status spread throughout the 13-month study period. At the end of the study, participants received a $100 honorarium and were permitted to keep their Fitbit devices.

### Variables

#### Participant characteristics and vacation status

At baseline, participants provided self-reported information on their sex, date of birth, relationship status, number of adults and children living in the household, smoking status, country of birth, Aboriginal and Torres Strait Islander peoples status, and any existing chronic health conditions. They also reported their highest level of education, total gross household income, employment status, and occupation. Baseline height and weight were obtained at the home visit. During the study, participants reported any dates of vacation where they were away from their usual residence. These days were excluded from analysis as local weather data was not available.

#### Movement behaviours

Minute-by-minute movement behaviour data were captured using the Fitbit Charge 3, worn continuously on the non-dominant wrist throughout the study period, except during charging. Participants were instructed to sync their device at least every five days, to ensure remote data capture via the custom-built “Fitnesslink” platform (Portal Australia, Adelaide, Australia).

Each minute of recorded data was automatically classified by Fitbit’s proprietary algorithm as sleep, sedentary behaviour, light physical activity (LPA), moderate physical activity, vigorous physical activity, or non-wear time. Fitbit devices have demonstrated acceptable validity for key behaviours in prior studies: sleep detection (compared to polysomnography: Charge 2 sensitivity = 0.96, specificity = 0.61^27^ and total sleep time Flex r = 0.97^28^), sedentary behaviour (compared to ActivPAL: Charge 3 ICC = 0.94; 95% CI: 0.92–0.96^29^), and moderate-to-vigorous physical activity (MVPA; compared to Actigraph: Charge 2 ICC = 0.69; 95% CI: 0.35–0.87^30^; Flex r = 0.73^31^).

For each participant day, total daily minutes of sleep, sedentary time, LPA and MVPA were calculated. A day was considered valid if the participant had at least 18 hours of device wear time, including a recorded sleep period. Participants were included in the study if they contributed at least two valid days. To normalise each day to 1,440 minutes, missing minutes were imputed from each participant’s minute-of-the-day behaviour profile, allocating each missing minute across behaviours in proportion to how that same minute was classified on their other valid days.

#### Temperature

Daily maximum and minimum temperature (°C) data for metropolitan Adelaide were sourced via the Australian Data Archive for Meteorology.^32^ Single daily values were calculated as the mean of readings from five major weather stations distributed across the Adelaide metropolitan region: Adelaide (West Terrace/ngayirdapira), Adelaide airport, Mt Lofty, Noarlunga and Parafield.

#### Climate change scenarios

Intergovernmental Panel on Climate Change (IPCC) Sixth Assessment Report (AR6) outlines numerous future climate projections. For this study, five future temperature scenarios corresponding to approximate central estimates for each Shared Socioeconomic Pathways (SSPs) were used.^33^ These ranged from lower-to higher-emissions futures: +1.5 °C warming (SSP1-1.9 by 2100), +1.8 °C (SSP2-4.5 by 2050), +2.7 °C (SSP2-4.5 by 2100), +3.6 °C (SSP3-7.0 by 2100), +4.4 °C (approx. SSP5-8.5). Current estimated warming, +0.99 °C (2000-2020 period) was subtracted from these projections to represent residual future warming relative to present-day conditions.^34^

### Statistical analysis

All analyses and visualisations were conducted in MATLAB R2024a.

From daily activity records across the full observation period, we estimated the probability of observing a behaviour with duration *d* at a daily temperature *t*, denoted *P*_behaviour_ (*d,t*). These distributions were constructed using 5-minute duration bins and 0.5oC temperature bins, yielding empirical temperature specific probability mass functions for each movement behaviour.

Temperature bins without behaviour data were filled using duration-specific temperature trends. by fitting a second-degree polynomial to the observed probabilities within each duration bin, allowing interpolation between observed temperatures and extrapolation beyond the observed upper temperature range.

These temperature-conditioned distributions were used within a Monte-Carlo simulation to estimate daily time (*d*) spent in each behaviour across a whole calendar year. For each simulated day, the input temperature (*t*), was the observed daily maximum temperature (daily minimum for sleep) plus a warming offset (relative to present-day temperatures) corresponding to one of the five IPCC climate scenarios.

Because MVPA exhibited a strongly zero-inflated distribution, it required an additional preceding step to account for its zero-inflated distribution, which involved estimating the probability of any MVPA within each 0.5oC temperature bin. Probability of any MVPA under simulated baseline conditions (i.e. 0°C warming) was calibrated to match the observed prevalence of MVPA days in the dataset.

For each climate scenario, daily behaviour was simulated across 1,000 iterations to estimate individual-level random effect. Results were aggregated by day-of-year to estimate population-level effects under each warming scenario. Outputs were summarised as empirically derived temperature-conditioned probability mass functions of daily behaviour duration and as temperature-ordered simulations quantifying differences in mean behaviour duration under future warming scenarios relative to the current-climate baseline (i.e. +0.99oC warming).

This framework operates at the population level, without individual-level random effects or covariate adjustment, to represent average temperature-behaviour relationships derived from all valid days in the dataset. These bin-specific distributions were used to simulate daily behaviour under the five shifted climate scenarios.

## Results

Empirical data from 85,182 valid days were available across 368 participants. The median number of valid days contributed per participant was 261 days (IQR = 179-343 days). The distribution of valid person-days across minimum and maximum temperatures is shown in Supplementary Figure 1.

Participant characteristics are presented in Table 1. The average age of participants was 40 years (SD = 5.9 years), 57% were female, and the majority were classified as overweight or obese (34% and 35% respectively). 85% were married or in de facto relationships and 90% had two or more children. Almost half the participants had a university education (48%), with 60% having an annual household income of $100,000 or more.

**Table 1:**
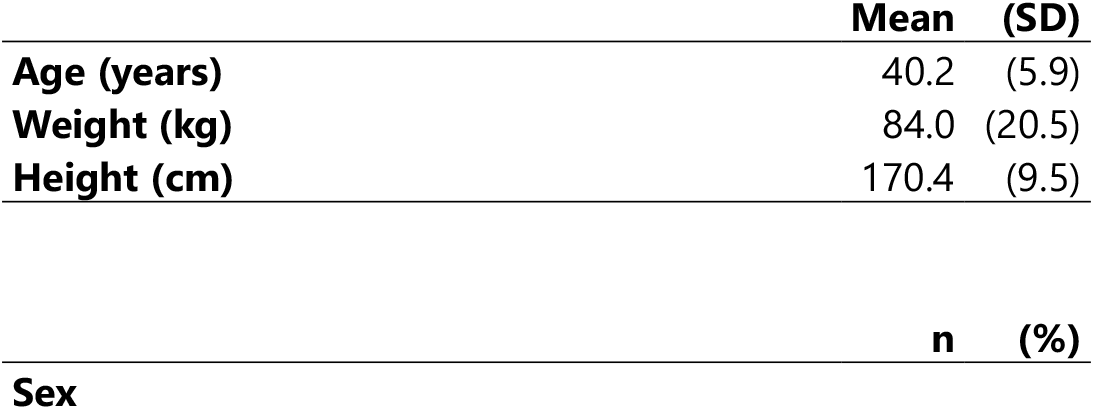
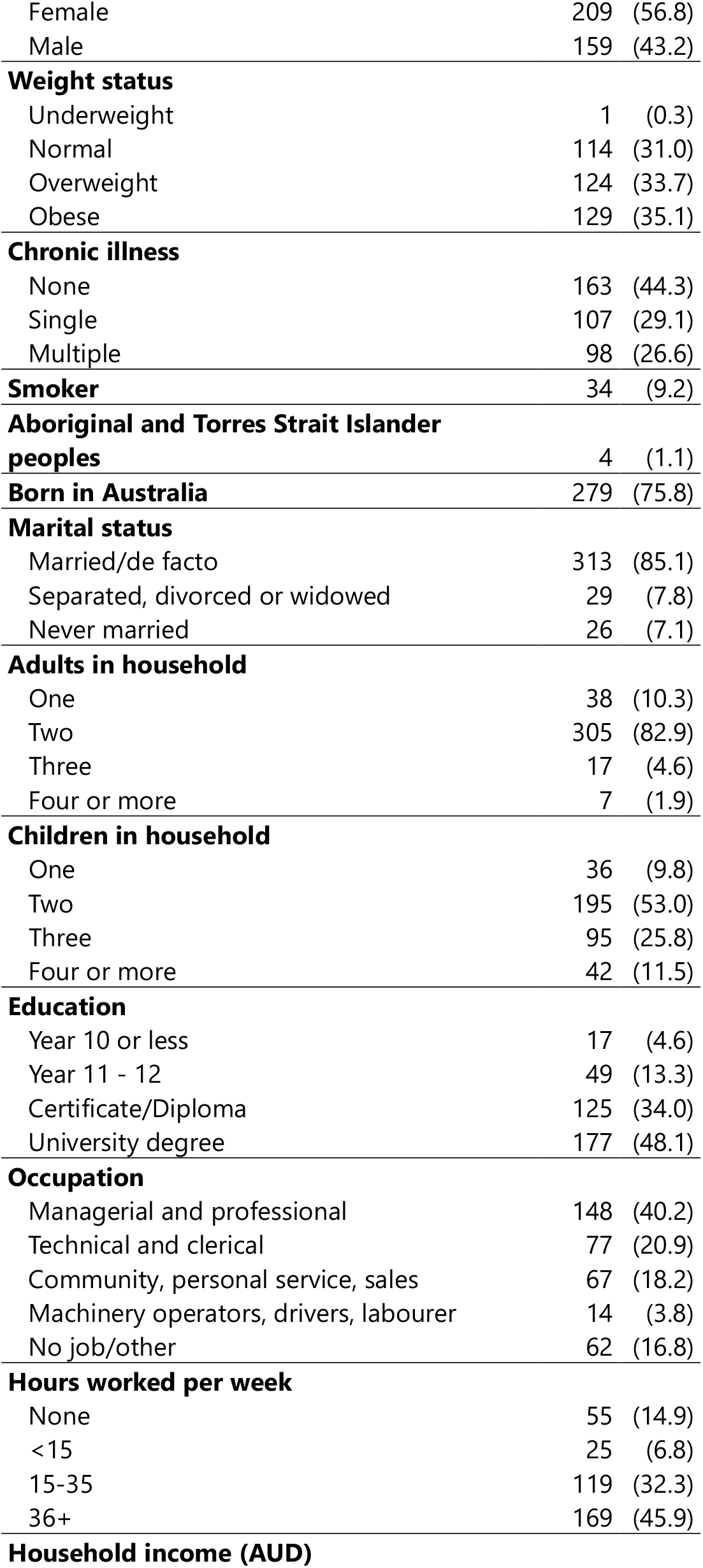
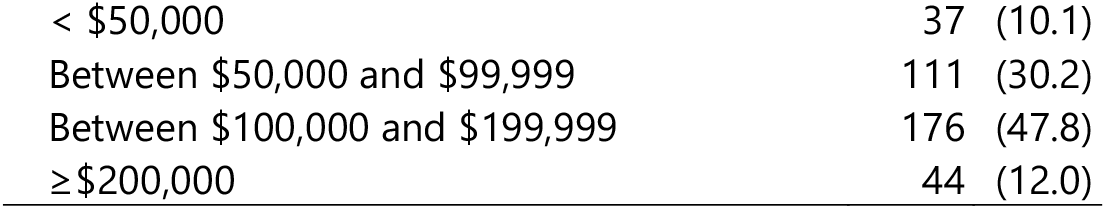
Participant characteristics at baseline (N=368)

### Distribution of daily movement behaviour durations

Figure 1 shows how the likelihood of spending different amounts of time in each movement behaviour varies with daily temperature. Rather than showing average values, the figure illustrates the full range of daily sleep, sedentary time and physical activity observed at each temperature.

**Figure 1:**
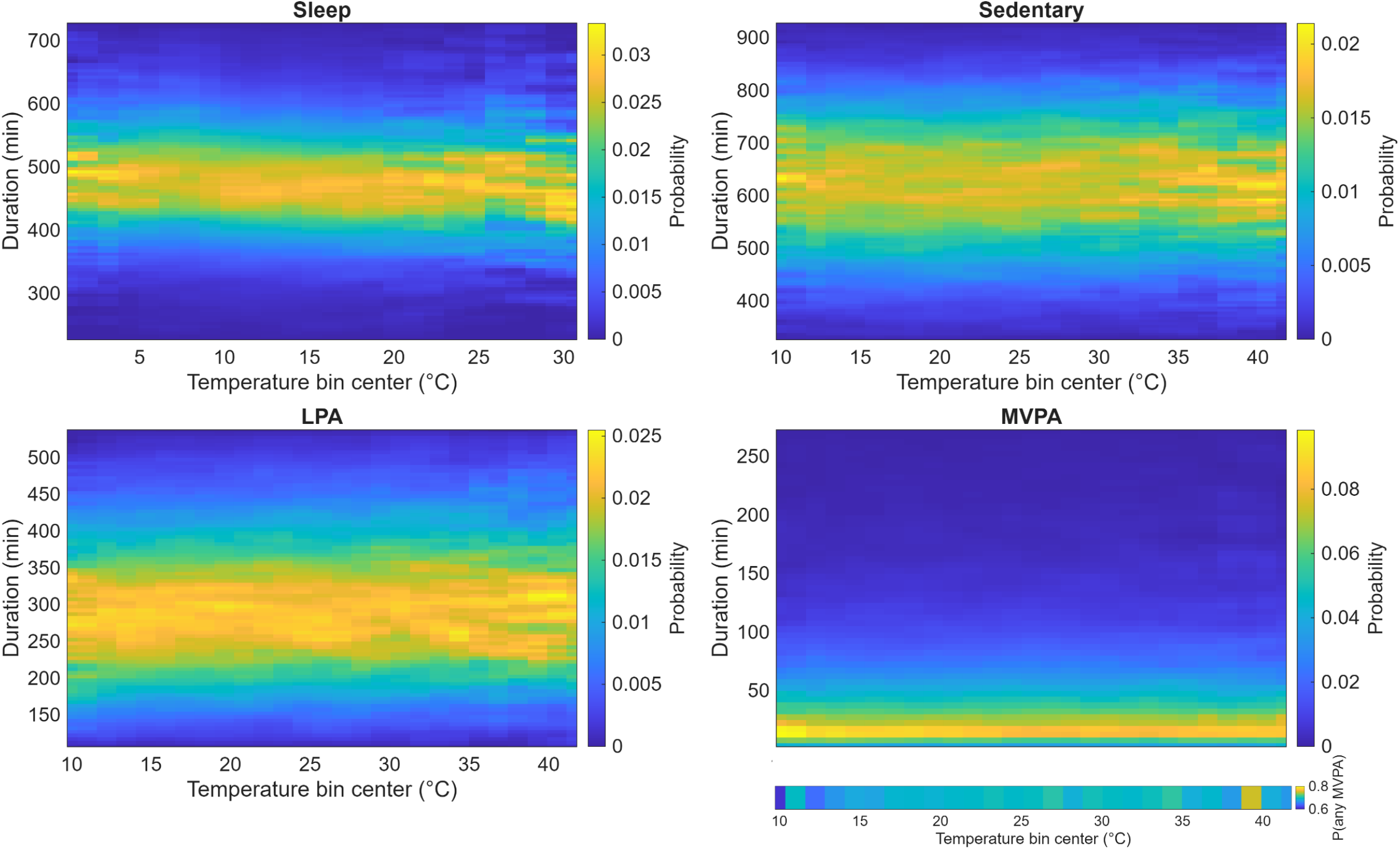
Probability mass functions (PMFs) of daily movement behaviour duration across 1oC temperature bins. Notes: LPA = light physical activity, MVPA = moderate-to-vigorous physical activity. MVPA also includes a PMF for “any MVPA”.

For sleep, warmer minimum temperatures were associated with a higher likelihood of shorter sleep durations, indicating that warm nights make short sleep more common. In contrast, sedentary behaviour showed little change across temperatures, with a wide range of daily sedentary time observed at all temperatures.

LPA also showed relatively stable daily patterns across the temperature range. MBPA was characterised by very low daily durations on most days at all temperatures, indicating that temperature mainly affects whether any MVPA occurs, rather than how much is accumulated on active days.

### Movement behaviour changes from baseline

Figure 2 shows how daily movement behaviours are expected to change under future warming scenarios, compared with current climate conditions, across the observed temperature range. Rather than showing average changes across the year, the figure illustrates how warming affects behaviour differently depending on the baseline temperature.

**Figure 2:**
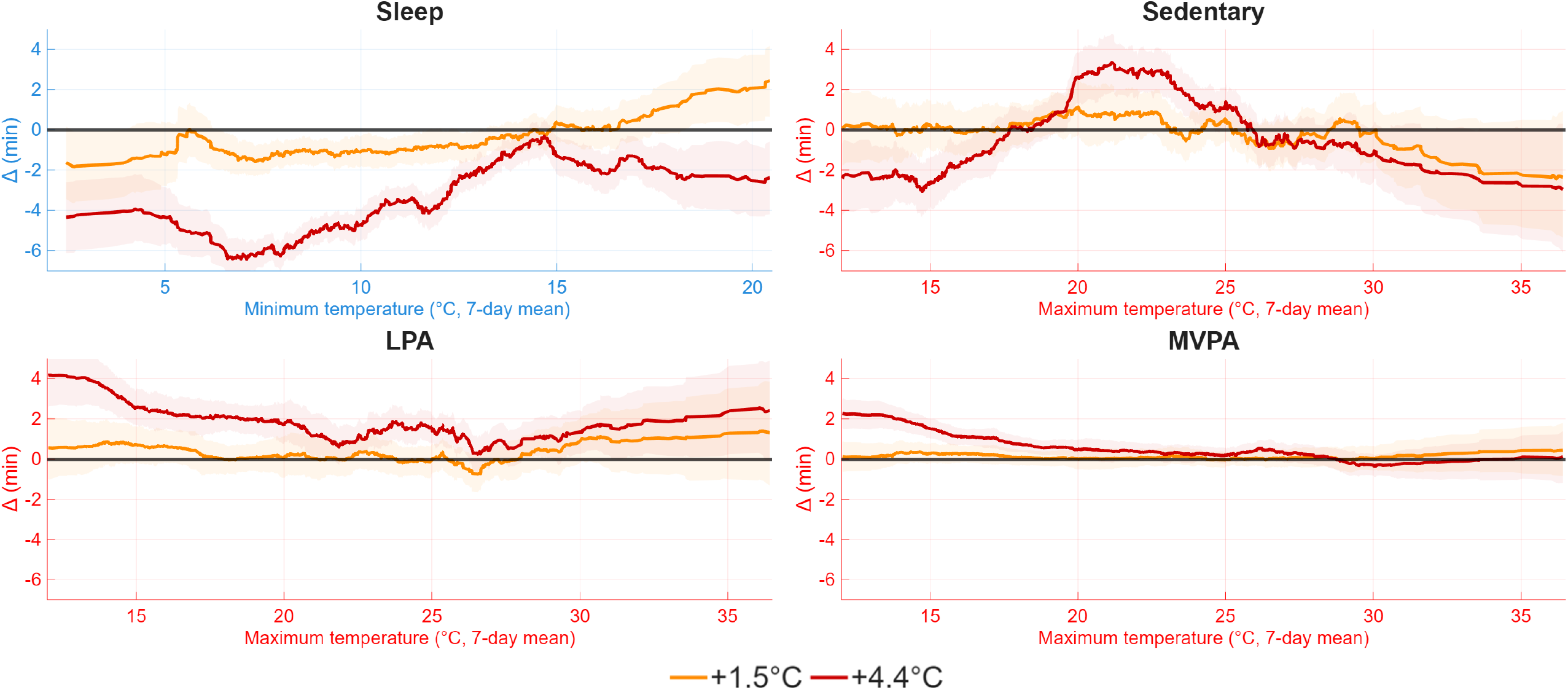
Temperature-ordered differences from current day movement behaviour durations under future warming scenarios. Note: LPA = light physical activity, MVPA = moderate-to-vigorous physical activity. Δ denotes the difference in simulated mean daily behaviour duration (minutes), calculated as scenario minus baseline. Baseline represents the current-climate scenario (+0.99°C warming from pre-industrial levels), and warming scenarios represent +1.5oC and +4.4oC relative to pre-industrial levels. Shaded bands indicate interquartile range across 1,000 simulation iterations.

For sleep, warming was associated with reduced sleep duration across most baseline temperatures. The largest sleep losses occurred on cooler days (approximately 5-10oC), with smaller reductions on warmer days. Across the temperature range, sleep losses were consistently greater under the high-warming scenario (SSP5) than under the low-warming scenario (SSP1).

Sedentary behaviour showed a more complex, non-linear pattern. Under warming, sedentary time tended to increase at moderate baseline temperatures (18-24oC), while decreasing at both cooler and hotter temperatures, particularly under SSP5. In contrast, LPA generally increased under warming across much of the temperature range, with the largest increases occurring on cooler days and a smaller dip at mid-range temperatures between 22-27oC.

Changes in MVPA were small overall. Under SSP1, MVPA showed little deviation from baseline across temperatures. Under SSP5, modest increases were observed, primarily on cooler days (<20oC).

Taken together, these patterns indicate that future warming is likely to redistribute movement behaviours across the temperature range rather than producing uniform increases or decreases, with sleep showing the most consistent sensitivity to warming.

### Year-long projections

Supplementary Figure 2 shows baseline, low- (SSP1) and high-warming (SSP5) scenario simulations across the calendar year. Overall, future warming was associated with modest but consistent changes in yearly average movement behaviours.

On average, time spent in physical activity increased under warming scenarios. Annual MVPA increased by a median of 49min per person (IQR= -3min to 1h 38min) under SSP1 and by 4h 22min (IQR= 3h 25min to 5h 16min) under SSP5, while LPA increased by 3h (SSP1, IQR= 1h 15min to 4h 56min) and 13h 01min (SSP5, IQR= 11h 16min to 14h 43min) per year relative to baseline. For MVPA, these increases were concentrated in the cooler months, with smaller reductions during the hottest periods. LPA was consistently higher than baseline levels under SSP5 across the year.

Changes in sedentary behaviour were small and inconsistent across scenarios and did not follow a clear seasonal pattern. Sleep duration showed the most consistent response to warming. Median annual sleep loss was -5h 29min (IQR= -7h 5min to -4h 6min) under SSP1 and -23h 19min (IQR= - 24h 53min to -21h 44min) under SSP5. Under SSP5, sleep was reduced in all months of the year, with the largest losses occurring during cooler months.

## Discussion

This study provides the first empirically grounded projections of all daily 24-hour movement behaviours under future climate scenarios. The results indicate that in a temperate zone with Mediterranean-style climate an increase in average daily temperature might result in small increases in time spent in MVPA and LPA in the cooler months, consistent decreases in sleep duration across the year and inconsistent changes in sedentary behaviour. These projections suggest that climate change in temperate zones may not negatively impact physical activity behaviour and might even increase physical activity modestly in the coldest months. By contrast, the steady year-round reduction in sleep duration is concerning, given the strong link between insufficient sleep and adverse health outcomes. Although these individual-level changes may appear modest, when scaled to populations they represent substantial shifts in health risk and economic burden.

Sleep projections observed in this study are consistent with findings from global studies of commercial datasets. Minor et al^19^ projected approximately 50 hours of excess sleep loss per person per year under mid-range warming (∼2.4± 0.5°C, RCP4.5), while Lechat et al^20^ and Li et al^21^ similarly reported reduced sleep duration with increasing temperatures. Although the magnitude of projected sleep loss in the present temperate-climate cohort was smaller, the direction and seasonal pattern were comparable. This convergence strengthens confidence in the temperature-sleep relationship and in the empirical modelling framework applied here, which differs from prior studies relying primarily on consumer-scale global datasets.

Projected changes in physical activity were modest in magnitude but directionally informative. Rather than a uniform decline under warming, the simulations suggest that physical activity responses are highly context-dependent, specifically seasonal patterns. In temperate climates, moderate warming may increase the number of days that are conducive to physical activity, without eliminating the constraining effects of extreme heat. This suggests that a redistribution of opportunity rather than a simple gain or loss.

Taken together, rather than indicating a simple linear response to rising temperatures, these patterns suggest a redistribution of movement behaviours across the temperature range and calendar year. In temperate climates, warming appears to shift a greater proportion of days into a thermal range that supports movement, while simultaneously narrowing conditions under which adequate sleep is maintained. This divergence reinforces the importance of considering the full 24-hour movement profiles, as apparent gains in physical activity may coexist with less visible but potentially consequential losses in sleep.

### Strengths and limitations

This study draws on a unique, high-resolution wearable-derived dataset capturing minute-level movement behaviours continuously across a full annual cycle. The ARIA dataset includes repeated observations across all seasons, allowing temperature-behaviour relationships to be estimated across the full range of conditions experienced in daily life.

A key strength of this work is the integration of these empirical behaviour distributions with a simulation framework that enables climate projections to be translated into plausible future behavioural patterns. This approach is readily transferable to other longitudinal movement behaviour datasets and provides a practical template for linking climate scenarios with population-level behaviour change.

Generalisability of the findings is limited to temperate-zone Mediterranean climate areas. Data collection overlapped with the COVID-19 pandemic. Although South Australia experienced comparatively limited and short-lived public health restrictions, pandemic-related routine changes may have influenced observed behaviour-temperature relationships. The results must be interpreted with caution because the study simulated temperature shifts in isolation, without accounting for other climatic changes such as humidity and precipitation, or their compound effects. The simulations applied a uniform daily warming delta across the year, whereas climate change is expected to increase the frequency and intensity of extreme heat events.^35^ Behavioural responses during heatwaves, potentially underestimated by uniform warming projections, could further modify or amplify the behavioural shifts observed here. The current methodology is adaptable to incorporating non-linear warming and heatwave scenarios.

The simulation is only valid for adults. On the other hand, this study prospectively recruited participants across multiple different socio-economic groups, which is a strength compared to consumer-based samples.

### Implications and future directions

Even modest daily changes in movement behaviours may translate into substantial health and economic consequences at the population-level. Health burden modelling demonstrates that small shifts in sleep and physical activity accumulate into large changes in disability-adjusted life years (DALYs).^36-38^ When coupled with mid-range cost-per-DALY estimates, behavioural pathways represent a large but under-recognised dimension of climate-health burden.^39^ These findings underscore the need for policymakers to consider movement behaviour adaptation explicitly within climate resilience strategies.

Importantly, the temperature-behaviour relationships observed here are unlikely to be uniform across populations. Emerging evidence suggests that vulnerability to heat-related sleep disruption is socially patterned. A study using data from the All of Us research program demonstrated that sleep duration reductions associated with high temperatures were 10-70% stronger among female, Hispanic populations, individuals of lower socioeconomic status, and those with chronic health conditions.^40^ Similarly, a small study of residents from a marginalised Los Angeles community, each 5oC increase in indoor night-time temperature was associated with 23 fewer minutes of sleep, suggesting substantially larger effects under conditions of limited cooling access.^41^ For physical activity, temperature influences leisure-time activity levels^42^, while neighbourhood design and access to green space shape opportunities for activity.^43^ Extreme temperatures are therefore likely to constrain outdoor activity and sleep more strongly in lower-resourced areas. These findings indicate that average population-level projections may mask important heterogeneity, and that climate-driven behavioural shifts are likely to intersect with existing social inequities.

These considerations highlight the need to better integrate climate science and behavioural epidemiology. Climate-adaptive movement behaviour epidemiology is an emerging area of research focused on understanding how projected warming alters 24-hour movement behaviours and how these changes may influence health. By applying defined climate scenarios to observed behaviour distributions, this approach enables estimation of how 24-hour movement patterns may evolve under different warming trajectories and how these shifts may vary across populations. Such projections can support the development of targeted adaptation strategies in public health and urban planning.

Future research should extend climate-scenario based modelling of 24-hour movement behaviours to diverse populations, particularly climate-vulnerable groups across a variety of climate zones. Incorporating non-linear warming trajectories, heatwave frequency, and additional environmental exposures such as humidity and precipitation will strengthen future analyses. Together, these directions establish a research agenda for climate-adaptive movement behaviour epidemiology and position behavioural forecasting as a critical component of climate-health planning.

## Conclusion

Climate change has the potential to subtly but meaningfully influence 24-hour movement behaviours in temperate Mediterranean climates. With rising temperatures, sleep duration declined consistently, while LPA increased and MVPA showed modest increases primarily during cooler periods. When projected across populations, these behavioural shifts have implications for future health burden. This study demonstrates how future climate projections can be integrated with 24-hour movement data to provide a foundation for incorporating behavioural pathways into climate-health modelling.

## Data Availability

All data produced in the present study are available upon reasonable request to the authors

## Acknowledgements

SM is a GenV Fellow and supported by a FAIR Fellowship 2024 Award administered by veski for the Victorian Health and Medical Research Workforce Action Plan on behalf of the Victorian Government. Funding for the Award has been provided by the Victorian Department of Jobs, Skills, Industry and Regions. Research at the Murdoch Children’s Research Institute is supported by the Victorian Government’s Operational Infrastructure Program. BL is supported by a National Health and Medical Research Council (NHMRC) of Australia Emerging Leadership Fellowship (2025886).

The authors would like to acknowledge colleagues who were involved in the design and conduct of the ARIA study, Emeritus Professor Timothy Olds, Emerita Professor Wendy Brown, Professor Adrian Esterman and Dr Gilly Hendrie.

## Author contribution statement

TF, SC, and CM conceived and designed the study. TF developed the methods with contributions from SC, CM, RC, SM, BL, and FF. TF performed the data analysis. TF drafted the manuscript, and all authors provided feedback, critical review, and approved the final version.

## Supplementary files

**Supplementary Figure 1:**
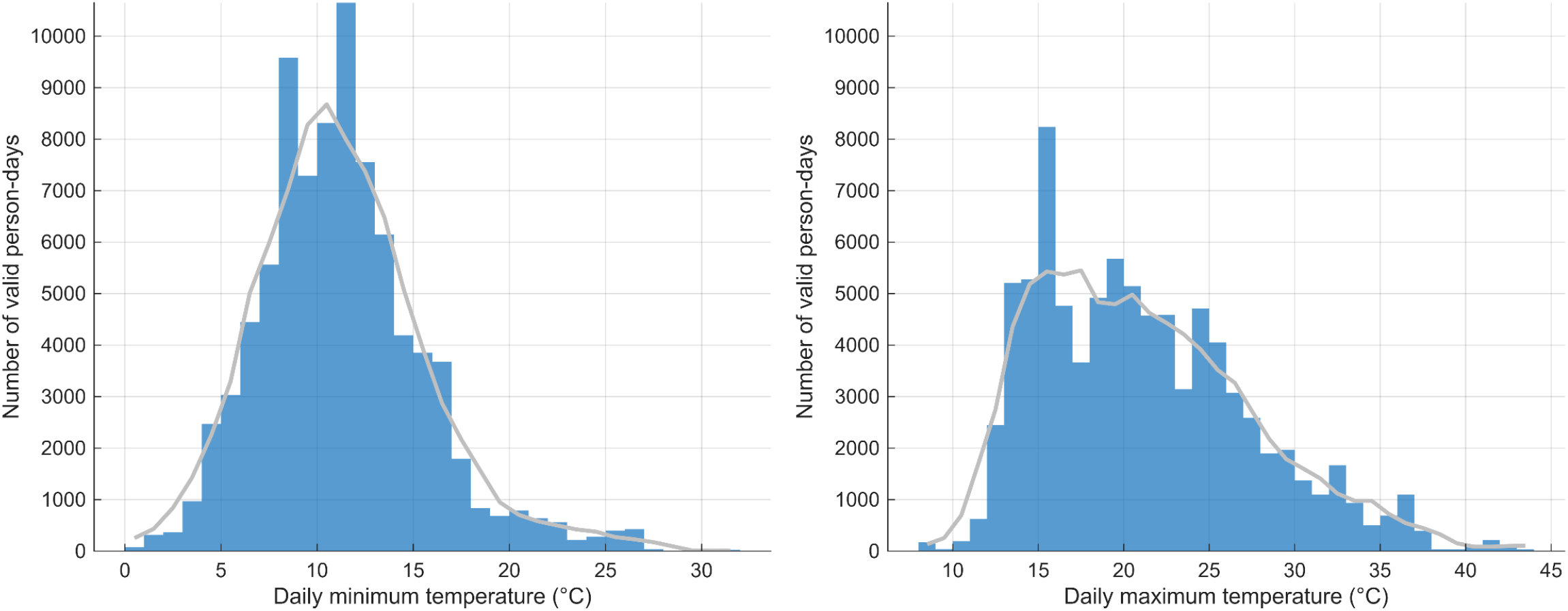
Distribution of valid person-days by minimum and maximum temperature

**Supplementary Figure 2:**
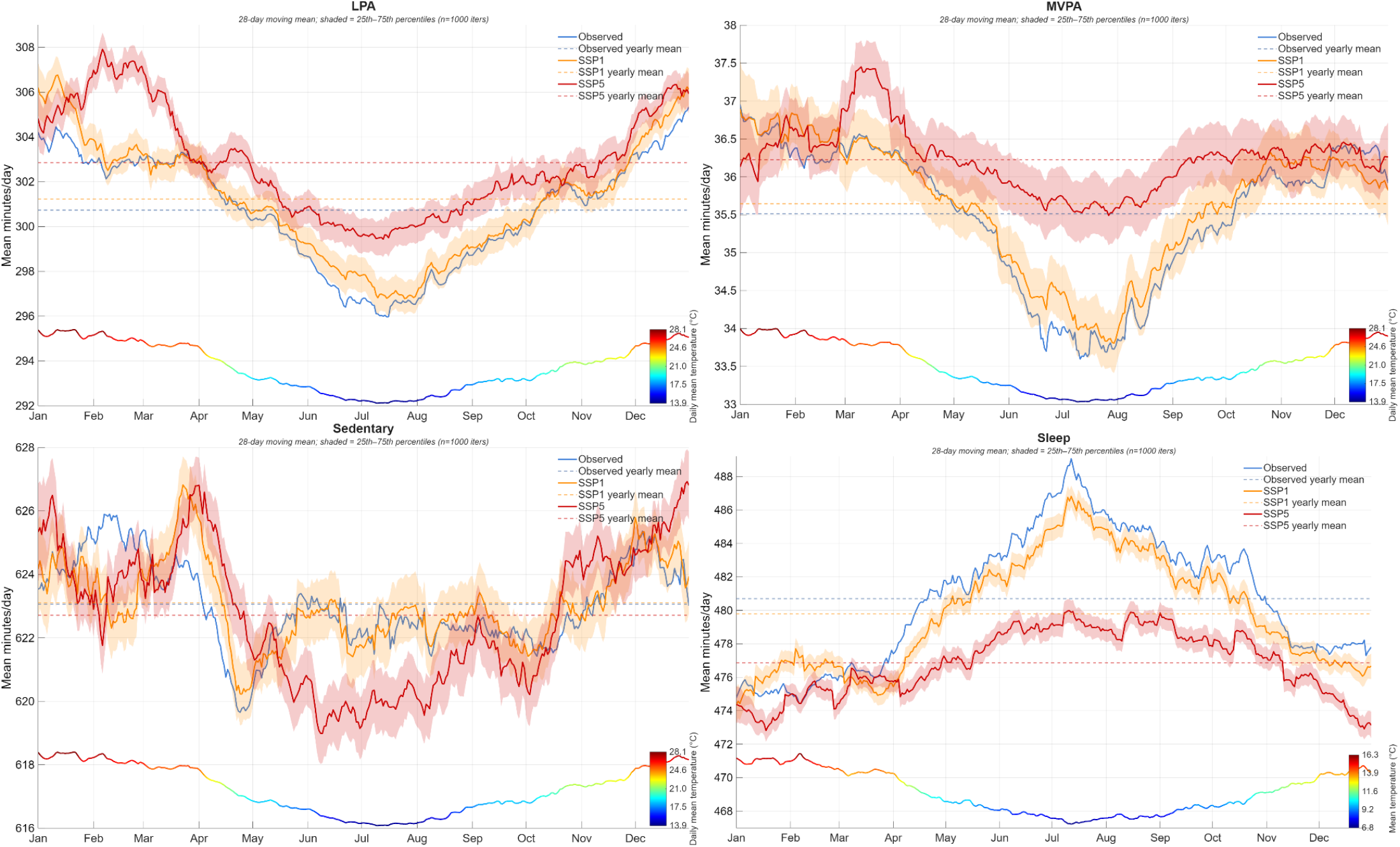
Movement behaviour modelling under observed, SSP1 and SSP5 temperature scenarios. Note: LPA = light physical activity, max = maximum, min = minimum, MVPA = moderate-to-vigorous physical activity, SSP = Shared Socioeconomic Pathway. SSP1-1.9 =+1.5oC warming over pre-industrial levels (residual of +0.51oC over observed data). SSP5-8.5 =+4.4oC warming over pre-industrial levels (residual of +3.41oC over observed data).

